## Supplementary Material for "ePOCT+ and the medAL-suite: Development of an electronic clinical decision support algorithm and digital platform for pediatric outpatients in low- and middle-income countries"

### Supplementary Information

### S1 Appendix: Prevalence of specific symptoms and diagnoses not covered in IMCI from Tanzania

**Table S1:** Notable symptoms and diagnoses for children above 2 months not existing in IMCI added to ePOCT+

| Additional non-IMCI conditions | Frequencies from Tanzanian studies/databases |
|---|---|
| ***Symptoms*** | |
| Headache | - 34% (0-5 years) and 81% (5-17 years) of febrile outpatients[1] |
| Abdominal pain | - 13% (0-5 years) and 23% (5-14 years) of febrile outpatients[1]<br>- 4.6% (2 months – 5 years) of febrile outpatients[2] |
| Sore throat | - 13% (5-14 years) of febrile outpatients[1] |
| Dental pain | - 30.2% dental pain among children age 12 to 19 years old[3] |
| ***Diagnoses / Classifications*** | |
| Fever without source (Undifferentiated febrile illness) | - 6.2% (0-5 years) and 7.1% (5-17 years) of outpatients[4] |
| Urinary tract infection | - 5.9% (2 months – 10 years) of febrile outpatients[5]<br>- 19% (0-5 years) and 16% (5-14 years) of febrile outpatients[1]<br>- 9.9% (0-5 years) and 19.7% (5-17 years) of outpatients[4]<br>- 8.1% (1 month – 5 years) of all outpatients[6]<br>- 18.6% (2-13 years) of febrile outpatients[7] |
| Eye disease | - 2% (1 month – 5 years) of all outpatients[6] |
| Trauma including burns | - 4.3% (0-5 years) and 21.6% (5-17 years) of outpatients[4]<br>- 9.7% (all ages) of outpatients[8] |
| Sexually Transmitted Infections | - 0.1 to 13.7% prevalence among adolescents 12 to 19 years old[9] |
| Dental caries and related oral problems | - 19.2% dental caries, 45.3% perceived need for dental care: prevalence among children age 13-19 years[3]<br>- 30.7% dental caries: prevalence among all age groups[10] |

### S2 Appendix: Delphi survey on the reliability and feasibility of measurement of symptoms and signs

#### Methods

A pre-selection of clinical elements were identified based on a systematic review on triage tools (ETAT, PEWS, pSATS, ESI, TOPRS, IMCI).[11] Symptoms and signs were excluded from the Delphi survey if a) the quality or predictive value was insufficient based on previous research [12], b) they are collected anyway during registration of the patient or c) they were known to be unfeasible for triage in Tanzanian primary care beforehand (for example laboratory tests not available at primary care health facilities). The Delphi survey was based on a recent Delphi study among international experts on predictors of sepsis in children under five[12] and included questions about each clinical element based on three domains: 1. Reliability of measurement, 2. Frequency of finding an abnormal value, and 3. Level of training required. Additionally, availability of instruments to measure vital signs and other challenges in collecting each element were evaluated. The answers were classified using a 5-point Likert scale: minimal, moderate, high, not applicable, I don't know. The answer options for the availability of vital sign instruments were yes/no/I don't know. We also collected data on the professional background and expertise of the participants.

#### Analysis

We analysed the results of the Delphi study according to the three domains. The answers were classified into a score 0-3: 0 = not applicable, and 1 – 3 for increasing strength of the answer. We calculated the total score of all participants per variable for each domain, resulting in a sum score of 0 - 90, stratified per level of care (dispensary or health centre). We also calculated the maximum score per variable, excluding the participants who answered ‘I don’t know’ to that particular variable. To facilitate comparison across items, we calculated the sum score as a percentage of the maximum score. We also created a composite sum score per item: sum score of domain 1 + score domain 2 – score domain 3, divided by the sum of max scores of all three domains, and separated for dispensaries and health centers. We did not predefine a threshold for inclusion in the final proposed triage tool. Analyses were performed in SPSS (version 25.0).

### Results

The results of the Delphi survey (number of participants=30) are shown in Table 2. While most signs and symptoms were feasible to assess at primary care health facilities, ‘capillary refill time’, ‘pain score (0-10)’, ‘assessment of cold skin’, and ‘weak and fast pulse’ had a lower score and were excluded from ePOCT+. Vital signs and anthropometric measurements including MUAC, heart rate and oxygen saturation also had a lower score due to lack of instruments and need for training.

**Table 2: Composite score on different domains per triage item**

| | **Combined score dispensary** | | | **Combined score health centre** | | |
|---|---|---|---|---|---|---|
| | *combined sum (domain 1+2-3)* | *combined max (domain 1+2+3)* | *%* | *combined sum (domain 1+2-3)* | *combined max (domain 1+2+3)* | *%* |
| General / past medical history* | | | | | | |
| Urgent referral status | 43 | 216 | 20% | 53 | 231 | 23% |
| Measurements / vital signs | | | | | | |
| MUAC (mm) | 7 | 195 | 4% | 16 | 207 | 8% |
| Temperature | 88 | 237 | 37% | 81 | 243 | 33% |
| Heart rate (HR) | 24 | 189 | 13% | 23 | 198 | 12% |
| Respiratory rate (RR) | 46 | 216 | 21% | 43 | 219 | 20% |
| Oxygen saturation (SpO2) | -10 | 159 | -6% | 7 | 174 | 4% |
| Pain score (0 - 10) | -4 | 150 | -3% | -7 | 153 | -5% |
| Weight | 80 | 234 | 34% | 73 | 234 | 31% |
| Capillary Refill Time | 10 | 162 | 6% | 12 | 180 | 7% |
| Airway / breathing | | | | | | |
| Central cyanosis / is the child blue? | 52 | 216 | 24% | 52 | 225 | 23% |
| Apnea (observed or reported) | 54 | 228 | 24% | 54 | 231 | 23% |
| Difficulty breathing (reported) | 73 | 240 | 30% | 77 | 249 | 31% |
| Difficulty breathing (observed: chest indrawing, grunting, nasal flaring) | 73 | 246 | 30% | 67 | 249 | 27% |
| Fast breathing (reported) | 79 | 240 | 33% | 75 | 243 | 31% |
| Circulation | | | | | | |
| Skin cold (cool peripheries) | 31 | 207 | 15% | 34 | 225 | 15% |
| Weak and fast pulse | 40 | 216 | 19% | 30 | 213 | 14% |
| Pallor - palmar, oral, conjunctival | 77 | 237 | 32% | 75 | 240 | 31% |
| Neurological | | | | | | |
| Irritability, restlessness | 87 | 237 | 37% | 84 | 240 | 35% |
| Convulsions (reported, history of) | 77 | 243 | 32% | 73 | 249 | 29% |
| Convulsing now, actively | 86 | 243 | 35% | 80 | 243 | 33% |
| Not able to drink or feed anything | 84 | 243 | 35% | 81 | 246 | 33% |
| Lethargy (AVPU) | 50 | 219 | 23% | 53 | 231 | 23% |
| Mobility - unable to move as normal | 41 | 222 | 18% | 37 | 222 | 17% |
| Dehydration | | | | | | |
| Sunken eyes | 77 | 243 | 32% | 68 | 240 | 28% |
| Reduced urine production | 47 | 210 | 22% | 43 | 210 | 20% |
| Infection | | | | | | |
| Fever (reported) | 103 | 234 | 44% | 95 | 240 | 40% |
| Gastrointestinal | | | | | | |

| | | | | | | |
|---|---|---|---|---|---|---|
| Diarrhea | 75 | 237 | 32% | 76 | 240 | 32% |
| Vomiting everything | 87 | 240 | 36% | 81 | 240 | 34% |
| Trauma | | | | | | |
| Significant trauma or other urgent surgical condition | 26 | 207 | 13% | 22 | 216 | 10% |
| Burns | 35 | 216 | 16% | 47 | 231 | 20% |
| Poisoning | 39 | 213 | 18% | 37 | 216 | 17% |
| Severe pain | 81 | 234 | 35% | 75 | 228 | 33% |
| **Average score overall** | | | **24%** | | | **23%** |

Footnote:
Colours = heat map per level of care, ranged from lowest % of maximum score to highest % of maximum score. Green = above average, red = below average, white = average.
*For duration of illness, number of previous admissions, admitted in past 2 days and history of HIV/sickle cell/palsy information was not available for all domains, so were left out of the composite score.

### S3 Appendix: Prognostic value of predictors used in the ePOCT and ALMANACH electronic clinical decision support algorithms

#### Methods

This is a sub-analysis of previously published data from the ePOCT study.[2] Briefly, a randomized controlled non-inferiority study was performed among children aged 2-59 months presenting with an acute febrile illness to 9 outpatient clinics in Dar es Salaam, Tanzania between December 2014 and February 2016. Patients were randomized by block to receive care using ALMANACH or ePOCT, a first- and a second-generation electronic clinical decision support algorithm (CDSA). The prognostic outcome for the present analysis was clinical failure by day 7. Symptoms, signs and tests were measured/assessed by study clinicians prompted by the respective eCDSAs.

#### Analysis

A bivariate logistic regression analysis and descriptive statistics were performed on a sample of clinically relevant predictors (table 3 and 4). Binary predictors were selected instead of continuous due to the binary cut-offs used within the IMCI chart booklet and ePOCT+ algorithm. Predictors with no observations within the two by two table were omitted from the analysis. Predictors for which the lower and upper 95% confidence intervals (CI) of the odds ratio (OR) do not overlap with 1, and the positive likelihood ratio (PLR) is 5 or above, or the negative likelihood ratio (NLR) is 0.2 or below, were considered to be significant predictors.

To further understand the independent prognostic value of each predictor, a multivariate logistic regression model with LASSO penalty was performed. A multivariable logistic regression for the probability of clinical failure was fitted to the predictors included in the bivariate analysis for the ePOCT and ALMANACH data sets. A LASSO penalty was used for feature selection. LASSO favours sparse solutions by shrinking less important coefficients to zero according to a penalty, which is proportional to the sum of the absolute values of the coefficients. LASSO exclusions are marked as “LASSO-excluded” in table 3 and 4. Some features appear several times in the bivariate analysis but are binarised at different thresholds (e.g. hemoglobin). To select the appropriate threshold for the multivariable analysis, a sequential feature selection was used, where a model for each available threshold was computed and compared according to the pseudo r-squared measures. The threshold that resulted in the best model was selected for inclusion into the final model. Excluded thresholds are marked as “redundant thresholds” in table 3 and 4. The odds ratio (with 95% confidence interval) was calculated using a multivariable logistic regression model without penalty for each remaining predictor (It is not possible to use the coefficients of the regression with penalty).

The per protocol population was used for this analysis since the intention to treat population considered all patients lost to follow-up as clinical failure. All analyses were performed using Stata (version 16) and Python 3.9.

#### Results and Discussion

Clinical failure at day 7 occurred in 2.3% (37/1586) of children managed using ePOCT, and 4.1% (65/1573) of children managed using ALMANACH. The bivariate logistic regression model found ePOCT danger signs (Unconscious, lethargic, 2 or more convulsions, or convulsing now) OR 12.4

(95% CI 2.5, 62); ALMANACH danger signs OR 13.5 (95% CI 6.6, 27.6), chest indrawing (ePOCT OR 9.3 (95% CI 2.9, 30.2); ALMANACH OR 17.2 (95% CI 7, 42.1)), hypoxemia <90% (ePOCT OR 29.4 (95% CI 4.8, 181.8)), respiratory distress (ALMANACH OR 7 (95% CI 3.2, 15.3) severe general appearance (ePOCT OR 9.4 (95% CI 2.8, 31.6), somnolence (ALMANACH OR 6.9 (95% CI 1.4, 34.1), any sign of anemia (ALMANACH OR 6.9 (95% CI 2.9, 16.6), and mid-upper arm circumference (MUAC) <12.5cm (ePOCT OR 12.3 (95% CI 3.3, 46.1); ALMANACH <11.5cm OR 53 (95% CI 12.9; 218.2)) to be prognostic of clinical failure. The multivariate logistic regression with LASSO penalty found respiratory distress and severe general appearance within ePOCT, and chest in-drawing, any sign of anemia and MUAC <12.5cm within ALMANACH to be prognostic of clinical failure. There are some limitations to the interpretation of this analysis. Notably, the prognostic value of each variable must be considered within the context of the original model. If the original model (ePOCT or ALMANACH) used a specific predictor to trigger a specific treatment or referral, then the prognostic value will likely be underestimated. This was apparent when comparing the prognostic value of very low weight-for-age z-score which resulted in antibiotics and a referral within the ePOCT algorithm, and not in ALMANACH.[13] As such, no rule was used to include or omit a clinical element within ePOCT+ based on this analysis, but helps contextualize how the algorithm branches can be improved. Future analyses however could specifically look at how the current algorithm can be improved in terms of prognostic and diagnostic accuracy, and model efficiency.

**Table 3: Bivariate and multivariate logistic regression model of clinical elements used in ePOCT to predict day 7 clinical failure**

| Prognostic factor | Bivariate analysis | | | | | Multivariate analysis with LASSO penalty |
|---|---|---|---|---|---|---|
| | OR (95% CI) | PLR | NLR | Sensitivity | Specificity | OR (95% CI) |
| **Binary variables** | | | | | | |
| Female | 1.1 (0.6, 2.1) | 1.0 | 1.0 | 46% | 56% | 1.3 (0.7, 2.6) |
| Hb <10 g/dL | 1.9 (0.9, 3.9) | 1.3 | 0.7 | 70% | 45% | 1.9 (0.9, 4.1) |
| Hb <7 g/dL | 1.9 (0.6, 6.4) | 1.8 | 1.0 | 8% | 96% | Redundant threshold |
| Hb <6 g/dL | 2.2 (0.3, 17.2) | 2.2 | 1.0 | 3% | 99% | Redundant threshold |
| Hb <5 g/dL | 4.8 (0.6, 38.5) | 4.7 | 1.0 | 3% | 99% | Redundant threshold |
| Chest indrawing | **9.3 (2.9, 30.2)** | **8.1** | 0.9 | 14% | 98% | 2.2 (0.5, 9.9) |
| Respiratory distress | **4.7 (2.2, 10.1)** | 2.9 | 0.6 | 50% | 82% | **5.0 (2.3, 11.0)** |
| ePOCT Danger signs (unconscious, lethargic, >=2 convulsions or convulsing now) | **12.4 (2.5, 62)** | **11.8** | 0.9 | 6% | 100% | Not kept in analysis |
| Diarrhea | 0.9 (0.3, 2.4) | 0.9 | 1.0 | 14% | 85% | LASSO-excluded |
| Very low weight for age (<-3 WAZ) | 1.5 (0.3, 6.3) | 1.4 | 1.0 | 5% | 96% | LASSO-excluded |
| MUAC <12.5cm | **12.3 (3.3, 46.1)** | **11.2** | 0.9 | 10% | 99% | 2.1 (0.6, 7.9) |
| Hypoxemia <90% | **29.4 (4.8, 181.8)** | **27.9** | 0.9 | 5% | 100% | 7.0 (0.8, 59.7) |
| Hypoxemia <93% | 4.2 (0.9, 18.4) | 4.0 | 1.0 | 5% | 99% | Redundant threshold |
| Respiratory Rate >=50%ile | 1.5 (0.6, 3.8) | 1.1 | 0.7 | 86% | 19% | Redundant threshold |
| Respiratory Rate >=75%ile | 1.3 (0.7, 2.4) | 1.1 | 0.9 | 54% | 52% | Redundant threshold |
| Respiratory Rate >=90%ile | 1.6 (0.8, 3.1) | 1.4 | 0.9 | 32% | 76% | 1.3 (0.6, 2.8) |
| Respiratory Rate >=97%ile | 1.1 (0.3, 3.6) | 1.1 | 1.0 | 8% | 93% | Redundant threshold |
| Heart Rate >=50%ile | 0.8 (0.4, 1.9) | 0.9 | 1.0 | 22% | 75% | 0.7 (0.3, 1.6) |
| Heart Rate >=75%ile | 0.3 (0, 2.5) | 0.4 | 1.1 | 3% | 92% | Redundant threshold |

| | | | | | | |
|---|---|---|---|---|---|---|
| General appearance (Normal, severe) | **9.4 (2.8, 31.6)** | 83.7 | 0.9 | 5% | 100% | **127.8 (10.7, 1525.7)** |

CI Confidence Interval; Hb hemoglobin; MUAC mid-upper arm circumference; NLR Negative Likelihood ratio; OR Odds ratio; PLR Positive Likelihood ratio; %ile percentile

**Table 4: Bivariate and multivariate logistic regression model of clinical elements used in ALMANACH to predict day 7 clinical failure**

| Prognostic factor | Bivariate analysis | | | | | Multivariate analysis with LASSO penalty |
|---|---|---|---|---|---|---|
| | OR (95% CI) | PLR | NLR | Sensitivity | Specificity | OR (95% CI) |
| **Binary variables** | | | | | | |
| Female | 1.3 (0.8, 2.1) | 1.1 | 0.9 | 51% | 55% | 1.2 (0.7, 2.0) |
| Danger sign (History of convulsions, unable to drink, unconscious/lethargic, vomiting everything, jaundice, cyanosis, stiff neck, severe pallor, severe wasting) | **13.5 (6.6, 27.6)** | **11.0** | 0.8 | 20% | 98% | 2.8 (0.7, 11.3) |
| Any sign of dehydration | **4.2 (1.6, 11.2)** | 3.9 | 0.9 | 8% | 98% | 2.1 (0.6, 7.3) |
| Sunken eyes | 3.8 (0.6, 24.8) | 2.7 | 0.7 | 40% | 85% | LASSO-excluded |
| Chest indrawing | **17.2 (7, 42.1)** | **14.0** | 0.8 | 20% | 99% | **6.1 (1.2, 32.1)** |
| Respiratory distress | **7 (3.2, 15.3)** | **5.8** | 0.8 | 20% | 97% | LASSO-excluded |
| Somnolence | **6.9 (1.4, 34.1)** | **6.8** | 1.0 | 3% | 100% | LASSO-excluded |
| Diarrhea | 1.1 (0.6, 2.1) | 1.1 | 1.0 | 21% | 81% | LASSO-excluded |
| Very low weight for age (<-3 WAZ) | **3.7 (1.5, 9.1)** | 3.5 | 0.9 | 9% | 97% | 0.3 (0.1, 1.2) |
| Respiratory Rate >=50%ile | 1.3 (0.6, 2.6) | 1.0 | 0.8 | 85% | 19% | Redundant threshold |
| Respiratory Rate >=75%ile | 1.7 (1, 2.8) | 1.3 | 0.7 | 63% | 50% | 1.4 (0.8, 2.5) |
| Respiratory Rate >=90%ile | 1.6 (1, 2.7) | 1.4 | 0.9 | 35% | 75% | Redundant threshold |
| Respiratory Rate >=97%ile | 1.3 (0.6, 2.7) | 1.2 | 1.0 | 12% | 90% | Redundant threshold |
| Heart Rate >=50%ile | 1.5 (0.8, 2.6) | 1.3 | 0.9 | 27% | 79% | Redundant threshold |
| Heart Rate >=75%ile | 1.7 (0.7, 4.1) | 1.6 | 1.0 | 10% | 94% | 1.3 (0.5, 3.5) |
| Any otitis incl discharge | 0.8 (0.1, 6) | 0.8 | 1.0 | 2% | 98% | LASSO-excluded |
| Any skin infection incl severe | 0.5 (0.1, 2.3) | 0.6 | 1.0 | 3% | 95% | 0.4 (0.1, 2.1) |
| Any sign of anemia | **6.9 (2.9, 16.6)** | **6.3** | 0.9 | 11% | 98% | **3.6 (1.2, 10.7)** |
| MUAC <11.5cm | **53 (12.9, 218.2)** | **47.5** | 0.9 | 11% | 100% | Redundant threshold |
| MUAC <12.5cm | 2.5 (0.3, 20.1) | 2.5 | 1.0 | 2% | 97% | **12.6 (5.0, 31.4)** |

CI Confidence Interval; Hb hemoglobin; MUAC mid-upper arm circumference; NLR Negative Likelihood ratio; OR Odds ratio; PLR Positive Likelihood ratio; %ile percentile

### S4 Appendix: Features of the medAL-*creator* and medAL-*reader software as defined by a clinical-IT collaboration with end-user feedback*

| Programming eCDSA platform (medAL-*creator*) | |
|---|---|
| **Feature** | **Description / rationale / example** |
| Easy platform so that clinician can program and/or review the algorithm | Drag and drop interface, obvious connectors, no visible scripts |

| | |
|---|---|
| Allow the integration of Weighted and Boolean algorithms to reach a diagnosis | Boolean (and, or, not);<br>Weighted algorithm; Based on a score using weighted variables |
| Allow for inclusion of sub-algorithms in any algorithm (predefined syndromes) | Ease the maintenance (and reduce risks of errors) for predefined syndromes that appear in several algorithms |
| Allow a diagnosis to exclude another diagnosis | Severe or complicated diagnoses can exclude non-severe and uncomplicated diagnoses |
| Allow a management to exclude other managements | Ex. Guidance to refer a patient to the hospital for one diagnosis excludes 'no referral' from another diagnosis |
| Allow a drug to exclude another drug | Ex. A broad spectrum antibiotic could exclude another narrow spectrum antibiotic |
| Ability to make a variable/question mandatory or not mandatory to respond | Allowing users the ability to skip non-essential questions to speed up processes |
| Allow the use of reference tables for clinical signs | In order to calculate z-scores and percentiles |
| Allow for cross-referencing of variables | To compute the BMI based on the weight and height of the patient, for instance |
| Allow conditioning of variables within the decision tree algorithm and for individual variables using specific filters | Variables only appear based on previous responses based on the decision tree logic, and based on certain filters (complaint categories) |
| Allow for management of multiple versions of different algorithms | Each version can be deployed to different users |
| Generation of data dictionary | Allow for future integration with alternative variable nomenclature (SNOMED, CID) |
| Algorithm validation mechanisms | An automatic validation process identifies errors in decision tree logic before an algorithm can be deployed |
| Modification restrictions to deployed algorithms | Restrictions to modify decision logic for algorithms that are implemented and in use (only minor modifications possible). New versions, however can make modifications and deployed allowing for HCW to understand the changes made. |
| Automatic conversion of the algorithm into a machine-readable code | Transforming "human-readable" drag and drop decision tree into machine-readable code for execution on the medAL-*reader* application |
| **eCDSA platform (medAL-*reader*)** | |
| **Feature** | **Description / rationale / example** |
| Multi-modal use on local network | Allow the use of different users, on different devices to manage a single patient |
| Ability for clinician using eCDSA to perform multiple, simultaneous consultations, with pause and resume capability | Allowing a health care provider to see another patient while sending another patient for laboratory investigations |
| Ability to accept, refuse, and add diagnoses and treatments proposed by the algorithm | To improve algorithms, monitor quality of care, and provide dosing for drugs not proposed by the algorithm. |
| To follow natural flow of consultation | First excluding emergency signs, evaluating the chief complaints, medical history, physical exam, investigations (laboratory tests), Diagnosis, treatment and management. |
| Option for user to move forward and backwards through the consultation process | To be able to update information from other stages if they are provided at a later stage |
| Follow-up questions/variables conditioned by root variables | Ex. Duration of cough, only to appear if cough present |
| Access to emergency management via an emergency button at any point during the consultation, even if for a different patient. | Without an emergency button to be able to press at any moment, a clinician will not be given immediate guidance for emergencies. |
| Alerts when clinician selects an emergency or danger sign | To be able to provide emergency management guidance if needed |
| Outline variables that would result in a referral | To motivate clinicians to assess danger signs with additional precaution |
| Division of medical history questions and physical exam signs by system | Organize consultation flow as clinicians are trained to; ie by system |
| Warning and error limit messages should advise clinicians of continuous values that are out of normal range, and out of feasible range. | To assure safety and quality of data inputted. |
| Provide option to give information and photos about each variable, diagnosis, drug and management. | Info buttons are placed beside each variable, diagnosis and management so that the clinician can get more information.<br>The images will be notably helpful for helping diagnose skin rashes and other physical signs. |
| Ability for user to state that some medical history questions are "unknown", some physical exams or anthropometric measurements are "not feasible" to measure, and some tests are not available. | In order to prevent the user from being blocked from continuing the assessment and prevent the input of false data. |
| Alert for user if they have not answered an important/mandatory question | To assure that clinically relevant and important questions are answered |
| A case summary is provided at the end of the consultation with the most urgent and important diagnoses listed first | To have a short summary of the previous consultation when the patient comes back for a follow-up or new visit |
| Ability to retrieve patient information from previous consultations using patient registration information | To facilitate follow-up consultations |
| Calculate medication dosing according to weight, age and formulation | Reduce error in medication dosing. |
| Support for translation | Allow for use of the same clinical algorithm in different languages |
| Have online and offline capacity | Health care workers should be able to use the tool online/offline |

| | |
|---|---|
| Data collection and synchronization to a central server | Through secure (encrypted) channels |
| Destination of the data from the app must be configurable | In order to comply with national regulators |

### S5 Appendix: Evaluation of ePOCT+ based on the characteristics set by the target product profile for electronic clinical decision support algorithm as defined by expert consensus[14]

| **General scope** | | |
|---|---|---|
| **Characteristics** | **Minimal / Optimal requirements** | **ePOCT+ / medAL-reader** |
| Intended use | Optimal | Captures diagnostic test results, patient clinical data to provide treatment and care recommendations |
| Target population | Optimal | Defined target population. Inclusion and exclusion criteria used when enrolling the patient |
| Setting | Optimal | Different algorithms used for different countries |
| Targeted end user | Optimal | Algorithms designed for use by nurses, physician assistants, but can also be used by medical doctors |
| Algorithm access | Optimal | medAL-*reader* app can be downloaded on android based devices |
| Algorithm content | Optimal and Planned | Based on WHO/international/local clinical care guidelines, peer-reviewed articles, and clinical experience/practice and clinical validation research.[2, 13, 15-18] For new algorithms clinical validation is planned in the form of cluster randomized trials. |
| Algorithm treatment recommendations | Optimal | Treatment recommendation based on international and national treatment guidelines, prioritizing medications available at the lowest level of care. Dosing calculated for clinicians. Treatment recommendations support antimicrobial stewardship. |
| Compatible POC tools | Optimal | POC tools used in routine care, and emerging diagnostic tools and devices relevant to the algorithm are included (CRP, Pulse oximetry) |
| Regulated toolkit components | Minimal | POC diagnostic tests and medical devices are regulatory approved, compliant with local regulations, and in the case of Tanzania included in the Standard medical laboratory equipment to be used at dispensary and health centre level (CRP, pulse oximetry, hemoglobin, HIV, malaria, syphilis, glucose)[19] |
| Compatible devices | Optimal | App is compatible with large smartphones, and tablets. (Compatible with computers through tablet mirroring) |
| Compatible operating systems | Optimal | medAL-*reader* compatible with android devices |
| **Clinical decision support algorithm** | | |
| **Characteristics** | **Minimal / Optimal requirements** | **ePOCT+ / medAL-reader** |
| Content transparency | Optimal | The healthcare programme and end user have access to underlying evidence and methodology used to develop the algorithm |
| Quality control | Optimal | The algorithm underwent both analytical and semantical verification. |
| Algorithm validation | Planned | While many of the algorithms have previously been validated, new content will be validated through a cluster randomized controlled trial |
| Machine learning | Planned | Machine learning models were used to help validated the use of certain predictors in the algorithms.<br>Following validation of the static ePOCT+ algorithms, machine learning models will be used to improve the algorithms, and validated in randomized controlled trials |
| POC data input | Optimal | POC data can be inputted in ePOCT+ / medAL-reader |
| Disease likelihood (POC tool) | Optimal | Prognostic positive/negative likelihood ration and pretest probability evaluated for all POC predictors (including Hemoglobin, glucose, pulse oximetry) in children presenting with fever from the community.[20]<br>CRP: Based on Diagnostic positive/negative likelihood ratio and pretest probability from the setting of interest.[21] Also evaluated in randomized controlled trial.[15] |
| POC training | Optimal | Training was provided to all end-users for all new POC tests/tools not normally used in routine care |
| **App** | | |
| **Characteristics** | **Minimal / Optimal requirements** | **medAL-*creator* and medAL-*reader*** |
| System validation | Optimal* | - Valid clinical association *And* clinical validation: Supported by well-established or novel evidence.<br>*Cluster randomized trials will be conducted to assure validity for algorithms without established evidence.<br>- Analytical validation: Multiple pathways for all algorithms were tested to assure that inputted data is processed correctly into expected output |
| System access | Minimal | Data access protected by authentication and authorization. |
| Context configuration | Optimal | Translation possible, country preferences for the algorithm can be configurable |
| Customisation | Optimal | Algorithms can be modified using medAL-*creator* including to updates to the list of medicines and POCs. |

| | | |
|---|---|---|
| User access rights | Optimal | Roles can be assigned to provide different levels of data access |
| Expert support | No | Access to online/remote expert advice to assist in patient consultation is not possible |
| App training | Minimal | On-site training |
| Internet availability | Optimal | Works offline and can trigger alerts for synchronization |
| Clinical data entry | Minimal | Manual entry by the operator |
| Patient management recommendation | Minimal | Consultation data summarized and actionable recommendations provided. Interoperability with EMRs and HIS is planned. |
| Navigation | Optimal | Non-sequential: the user can move to a certain extent in any direction through an assessment and change input data to reach a final recommendation |
| Workflow requirements to enable time-delayed POC data input | Minimal | User can perform multiple, simultaneous consultations, with pause and resume capability, to allow clinical and laboratory data entry |
| Task management | Optimal | Multiple algorithms can be supported simultaneously in one application against a common data set |
| Follow-up | Optimal | Ability to retrieve patient information using patient registration information. However data from previous consultations cannot be automatically integrated within the algorithms for the new follow-up consultation |
| System malfunction protection | Optimal | System malfunctions are made clear to the user |
| Scalability | Optimal | The app allows for high transaction volumes with complex workflows to cover primary care workforce at a national scale |
| Updates and versioning | Optimal | Processes are in place to control any app changes (including algorithm version updates) and provide the appropriate and correct update to the user |
| **Data** | | |
| **Characteristics** | **Minimal / Optimal requirements** | **medAL-*reader*** |
| Data capture | Optimal* | Can capture text, image, numeric, GPS, barcode.<br>Does not capture audio, video or biometric |
| Data validation | Optimal | The warning and error alerts can be programmed to prevent errors of data input |
| Data ownership | Optimal | The healthcare programme of the country of implementation has ownership of the data |
| Data storage | Optimal | The healthcare programme can choose the destination of the app's data |
| Data recovery | Optimal | The system can be re-established to the desired state in the event of interruption or failure. Data is saved upon completion of each stage (registration, 1st assessment, medical history and physical exam, tests, and diagnosis and management.) |
| Data flow | Optimal | The flow of data is determined by the healthcare programme |
| Data reporting | Optimal | Dashboards will be configured to present real-time data for reporting, benchmarking and monitoring |
| Data provenance | Optimal | Provides origin and processes applied to output data. When data are downloaded or shared, the version of the model is tagged so it is always clear how the data was obtained |
| Data dictionary | Planned | Data dictionary is automatically created by medAL-*creator*. Link to international reference standard terminology in development. |
| Data security and privacy | Optimal | The app operates under secure connectivity which meets data protection and regulations of individual countries to avoid loss and corruption of sensitive data, and mitigate cyberattacks, whether data are at rest or in transmission.<br>Includes:<br>► Authorisation/access control<br>► De-identified data<br>► Data encryption<br>►Two-factor authentication |

CRP, C-reactive protein; GPS, Global Positioning System; POC, point-of-care; WHO, World Health Organization
